# Advancing Hair Loss Assessment in Alopecia Areata: The Mathematical Case for Centralised, Standardised Imaging

**DOI:** 10.64898/2026.04.02.26349939

**Authors:** David Fleet, Alyson Bryden, Andrew Messenger

**Affiliations:** Soterios Ltd, Laburnum House, East Grimstead, Salisbury, Wiltshire, UK; Departments of Dermatology, Ninewells Hospital/Medical School, Dundee, UK; Division of Clinical Medicine, University of Sheffield, UK

## Abstract

**Background:** In clinical trials for alopecia areata (AA) the treatment effect (percentage of hair loss) is estimated using the Severity of Alopecia Tool (SALT) score. Trials in patients with severe AA (≥50% hair loss) employed a “local” rating of the SALT score performed at trial sites by different investigators. However, in mild-to-moderate AA (≤50% hair loss) where SALT scores are lower, potential inter-rater variability and margin of error may compromise the results.

**Objectives:** To compare Centralised and Local measurement of hair loss in mild-moderate AA.

**Methods:** In a Phase 2 clinical trial a centralised measurement of hair loss was performed from photographic images taken using a standardised protocol and professional camera equipment. Local scoring was also undertaken at screening/baseline for eligibility. We assessed: the repeatability of the central system (screening vs baseline values), the reproducibility of the central versus the local rating system and the potential impact of each method on the endpoints using a Monte-Carlo simulation method.

**Results:** There was good agreement and consistency of scoring with Central rating. This provided much smaller margins of error, 50% lower than Local rating. The simulations demonstrated that substituting Local rating for Central rating would result in a reduction of the likelihood of a statistically significant outcome by at least 50% depending on the SALT score defined clinical response endpoint.

**Conclusions:** Central rating is most appropriate in the Phase 2 ‘learning’ stage of clinical development and provides an accurate representation of the quantity of hair loss, minimising error and ensuring consistency in measurements.

**‘What is already known about this topic?:**

- Hair loss in alopecia areata (AA) multicentre clinical trials is measured using the SALT score
- In severe AA (SALT 50-100) local ‘in-person’ assessment is recommended to monitor treatment effects but here margins of error for changes have a wide tolerance
- In mild/moderate AA (SALT<50), where smaller changes are likely, measurement accuracy and precision are essential
- Published standardised imaging processes supporting the use of Central rating lack the required numerical analysis support this report provides

**‘What does this study add?:**

- Local rating is not appropriate as scoring lacks accuracy and precision required in the mild/moderate condition
- The value of Central rating over Local assessment is confirmed

## Introduction

Research into the methods of measuring hair loss in alopecia areata (AA) revolve around the Severity of Alopecia Tool (SALT) score.^1^ The use of this tool is routinely accepted,^2^ although there is considerable debate as to how this score measurement is actually performed during assessment because of the debate over hair margins and sometimes difficulty with irregular or diffuse areas of hair loss.

In clinical trials involving the severe AA condition^3,4^ local rating was performed but, in these situations, large scale changes were expected. Patients were included with SALT scores around 100 and SALT score <20 were required to demonstrate improvement with treatment. On the face of it consistent scoring of the severe condition in clinical trials would appear easy to achieve. However, even here there was concern over the trial Investigator’s lack of familiarity with the measurement, and emphasis was placed on the use of photography in repeated training sessions with the local Investigators.^5^ In addition to practical issues, numerically consistent and accurate scoring has also been shown to be problematical, with scoring of the same images in severe AA by 8 trained dermatologists giving mean differences in the order of 10%.^6^ Interestingly this paper also advocated the use of integrated software (a neural network-based program) to further assist the dermatologists define hair margins and improve scoring, something not possible with local assessment.

Although there is no precedence for either local scoring or central rating as the preferred measurement in mild to moderate AA cases, there is evidence from other skin conditions, notably psoriasis using a similar outcome measure (the PASI).^7^ So much so, that it is stated that the margin of error is unacceptable at the lower end of the rating scale (PASI <20), with poor correlation (agreement) between raters, and that local scoring therefore should be ‘avoided at all costs’.

In our Phase 2 double-blind, placebo controlled clinical trial in mild to moderate AA patients (guideline SALT 10-50) a centralised measurement of hair loss was performed from photographic images taken using a strict protocol and standardised professional camera equipment. Local scoring was also undertaken at screening/baseline for eligibility. Here, we have used a numerical analysis approach to compare the centralised and local measurements of hair loss in mild-moderate AA. We assessed the repeatability of the Central system (screening vs baseline values), the reproducibility of the Central versus the Local rating system, and the potential impact of each method on the endpoints using a Monte-Carlo simulation method.

## Materials and methods

Central rating SALT scoring was performed by a single experienced rater using multiple photographic images taken by nursing staff after training and following a specific protocol using specifically configured camera equipment uploaded to a software application which randomly presented batches of photos and provided a moveable grid and zooming to aid visual scoring. The SALT scoring process performed (see SOT001 Clinical Trial Imaging Charter (Appendix)) was in line with approved guidelines.^8,9^

Local scoring was undertaken at Screening and/or Baseline (Visits 1/2) for the purposes of eligibility and enrolment. Although no standard local training was performed all such raters were experienced Dermatologists or were previously trained to perform SALT scoring. Central rating was performed at both Screening and Baseline visits to allow assessment of repeatability (consistency) and at all subsequent visits during treatment after 2 months, 4 months and 6 months (Visits 3, 4, 5) to accurately assess change over time. Comparison between Local and Central rating performed prior to treatment was also performed to assess reproducibility (comparison).

In addition, at each visit prior to and during treatment, along with imaging for central rating, a specific patch was selected and photographed ‘close-up’ and the area (mm^2^) accurately measured using commonly used available software (Image J). As a further assessment of the validity of Central scoring the two sets of results were compared after standardising (with values expressed as a proportion of the maximum observed score). Correlation between Central scores and the Patch area measurements was also performed to support Central scoring.

Statistical analysis of repeatability and reproducibility between methods with assessment of agreement, error, bias, variability and correlation was performed using standard methods.^10,11^

### Repeatability (Consistency)

The Central method performed on separate occasions at Screening and Baseline assessments were considered paired subject values. Agreement was quantified by the within-subject standard deviation-SD (sqrt(2)*wSD). to estimate the size of measurement error. The repeatability coefficient (1.96*sqrt(2)*wSD) was derived to estimate the variability of the agreement value (with 95% confidence interval). These data are presented in Bland-Altman scatter plots of the differences vs mean values for all subjects. The mean difference together with -/+ 1.96*SD(difference) are overlaid as lines of estimation.

An ANCOVA model was fitted to the repeated measurements (pairs of values) with both within (wSD) and between subject (bSD) variability determined to partition true or ‘error-free’ values of subjects and measurement error.

The reliability of the agreement estimates was also determined by calculating the intra-class correlation coefficient (ICC). The ICC, a measure of heterogeneity in the subject population, was defined as bSD**2/((bSD**2)+(wSD**2)). Larger values indicate that more of the variability is apportioned to the true subject values and less to the measurement error.

Further assessment of the accuracy of Central Rating with software-measured specific patch areas was performed using Pearson correlation statistic of the change from baseline values for the two methods after standardising the two as a proportion of the maximum change observed

### Reproducibility (Comparison)

Both the Central and Local method scores were measurements on the same scale, and to evaluate the relationship and potential bias between the two methods, one is plotted against the other with a line of equality (Central vs Local). Bias is evident if there are more data points above/below this line, showing that one method gave consistently higher values than the other method.

To quantify the differences between the measures a Bland-Altman plot was used, as in the repeatability evaluation with differences between measurements (Central vs Local method) and mean subject values. The difference from the zero provides an estimate of the bias between the two methods. The limits of agreement quantify the expected difference in the measurements and these are calculated as: mean difference -/+ 1.96*SD(difference). The limits of agreement assessment assume that variability (SD) of the method differences are uniform throughout the range of measurements. Association between the differences and the means was investigated with a regression plot in comparison with the line of uniformity

Under or over estimation of measurements (bias) was then investigated using a one sample t test of the mean differences, which tests the hypothesis that the true mean is zero and there is no bias. Confidence intervals (CIs) for the bias were calculated as mean difference -/+ 1.96*SE(difference) (SE=SD(differences)/sqrt(N)).

A linear regression analysis investigated the relationship between scores for the Central and Local method. Estimates of the slope together with ‘predicted’ standard errors (SE) are presented.

### Simulation model

A simulation model (Monte-Carlo method) using the relationship between Central and Local scores observed at baseline was used to assess the likely impact on the Phase 2 trial if Local scoring throughout was adopted. This involved determination of the distribution of the difference (Figure 1: approximately normal with mean: 2.7 and SD 13.6) with random selection of observations applied to each of the change from baseline central scores for the 155 individual patients, then derivation of the new local percentage change from baseline and the categorised Responder/Non-responder grouping (>30%/100%) repeated on up to 10,000 occasions to represent different trials with each one analysed separately using ANCOVA model and Fishers exact hypothesis test, respectively. The proportion of correctly predicted statistically significant outcomes (p<0.05) was then used to indicate the power of the tests (Type II error=probability of declaring no difference when a difference really exists)

**Figure 1.**
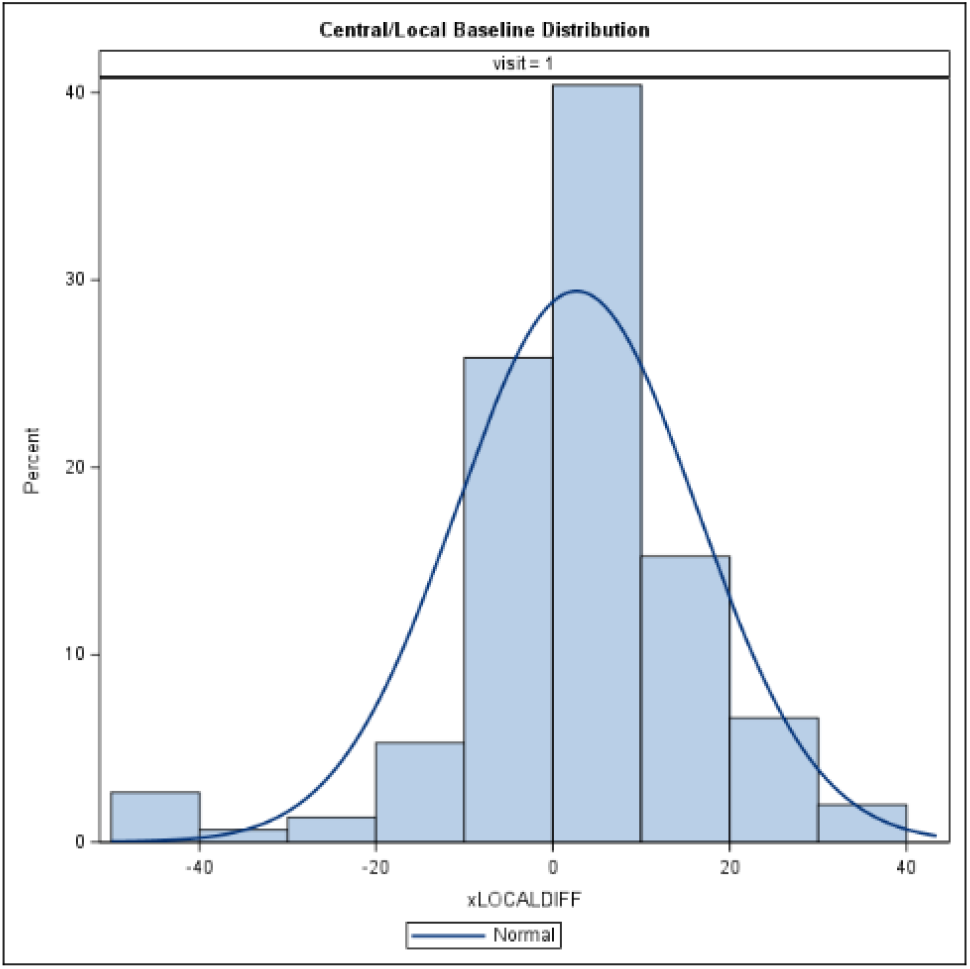
Central/Local Rater Distribution of SALT scores at Baseline.

## Results

Results are presented for all SALT scores together and separately for SALT scores <20/≥20 to help outline differences observed, particularly at the lower end of the scale

### Central Rating: Repeatability (Consistency)

Overall SALT scores Central-Central rating was extremely consistent with an Agreement (measurement error) of 5.43, repeatability coefficient of 10.6 and ICC of 0.954

The paired values from screening and baseline assessments were used to quantify the measurement error using Bland-Altman plots of differences vs means (Figure 2A). The plot includes lines representing the mean difference and the lower and upper Limits of Agreement (LoAs=10), given by the mean difference ±1.96 SD. The results show good agreement and consistency of scoring with Central Rating although there appears to be some increase in the variability when going beyond SALT 20. No bias is evident as mean values are centred on zero throughout.

**Figure 2A:**
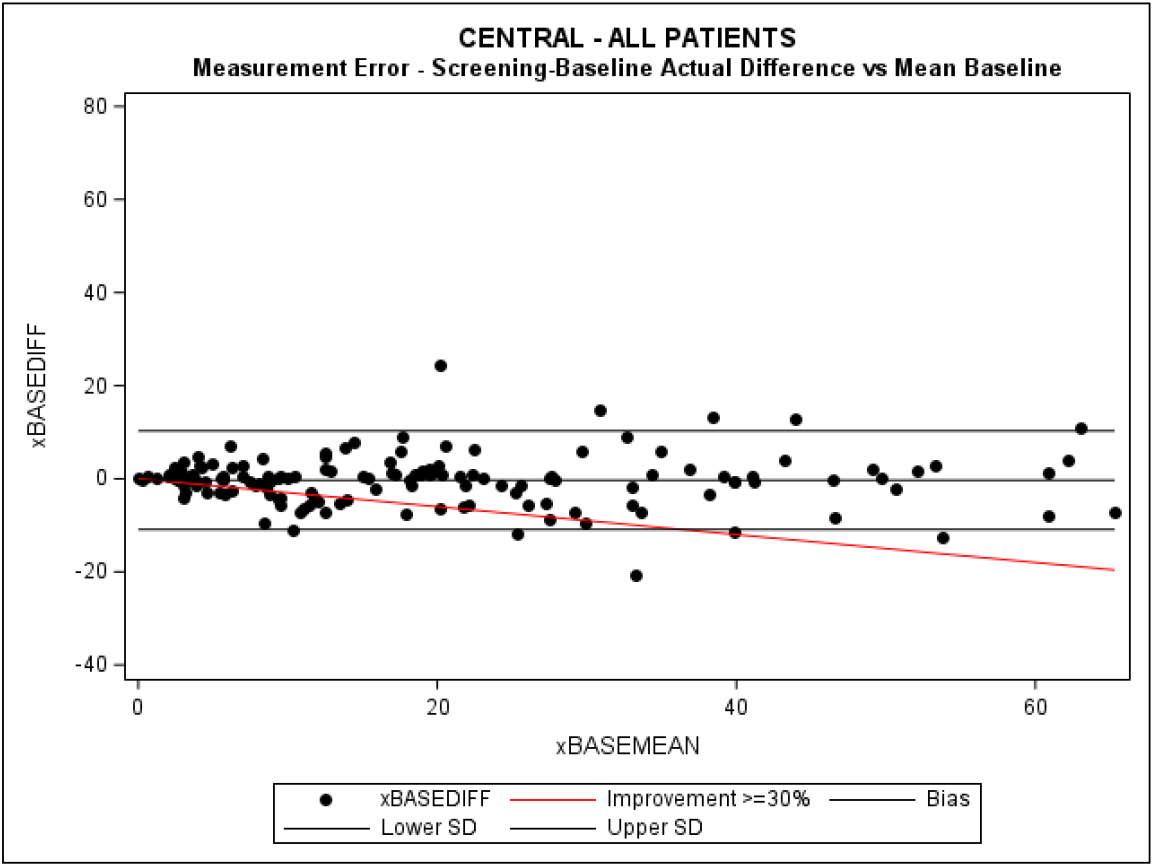
Repeatability (Consistency) – Central Rating Baseline vs Screening (All SALT Scores) NB: Red line indicates level to attain during subsequent treatment to achieve >=30% improvement

Separate plots for SALT<20 and SALT≥20 (Figure 2B) suggest LoAs of 5 and 15 for the two, respectively, suggesting the overall variability is inflated the higher the SALT score.

**Figure 2B:**
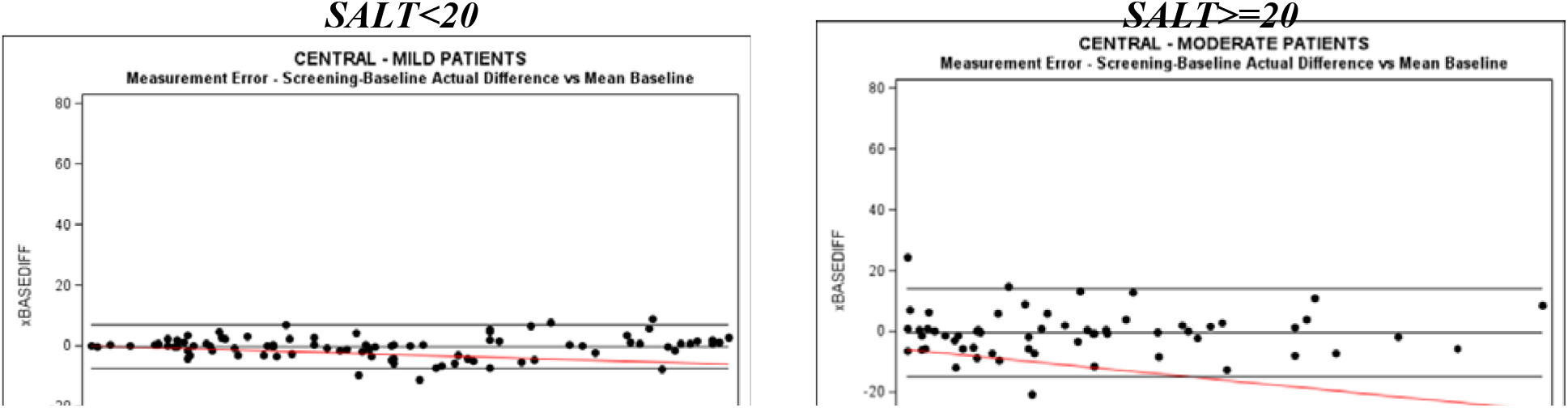
RRepeatability (Consistency) – Central Rating Baseline vs Screening by Group.

Correlation plots (Figure 2C) for baseline vs screening Centrally rated scores demonstrate excellent agreement and confirms minimal bias against the line of uniformity (black line).

**Figure 2C:**
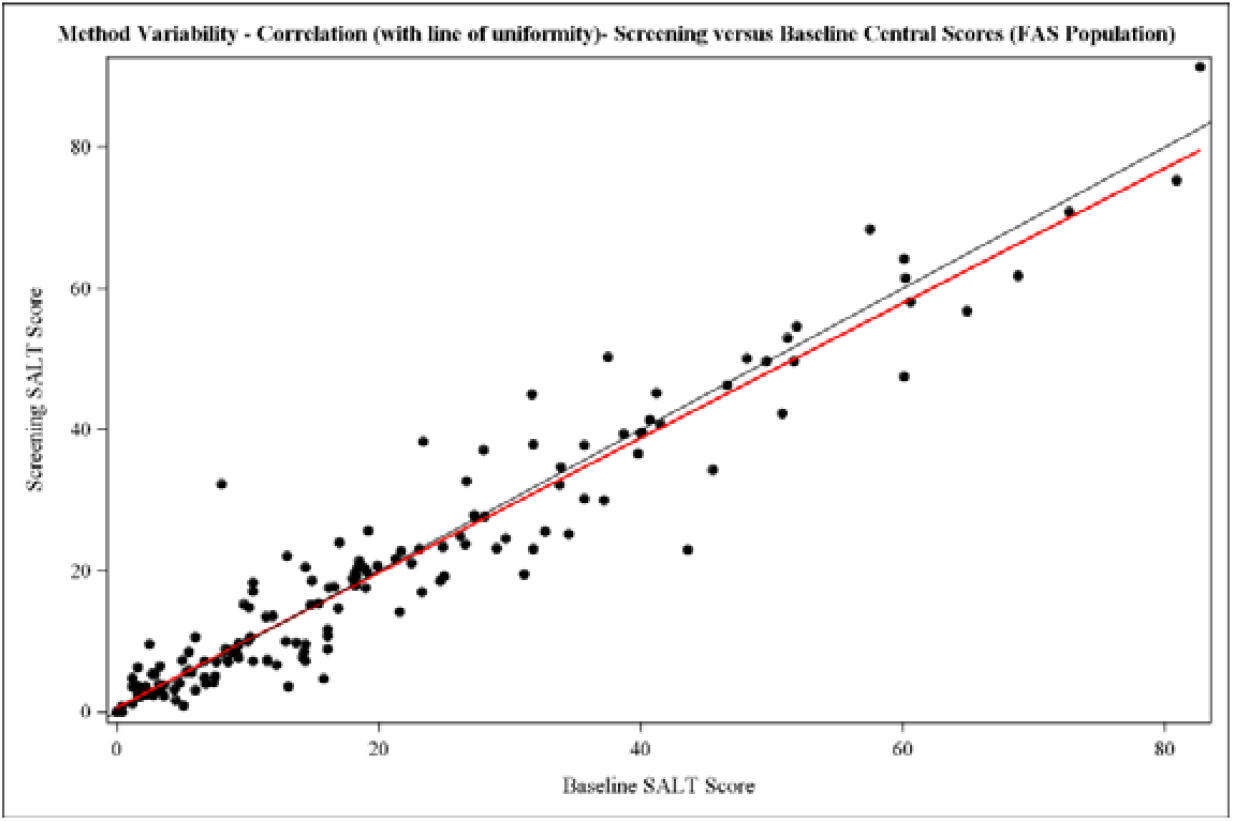
RRepeatability – Measurement Bias: Central Rating Baseline vs Screening.

### Central rating: Correlation with patch area measurement

Correlation plots across all SALT scores and SALT scores<20 (Figure 3) for the difference from baseline scores as measured for all visit assessments (from 2 months, 4 months and 6 months) by Central Rating vs Patch Area demonstrates very good agreement around the line of uniformity (albeit with two very different measuring procedures and objectives) with r coefficients of 0.40 across all SALT scores increasing to 0.48 with SALT scores <20.

**Figure 3.**
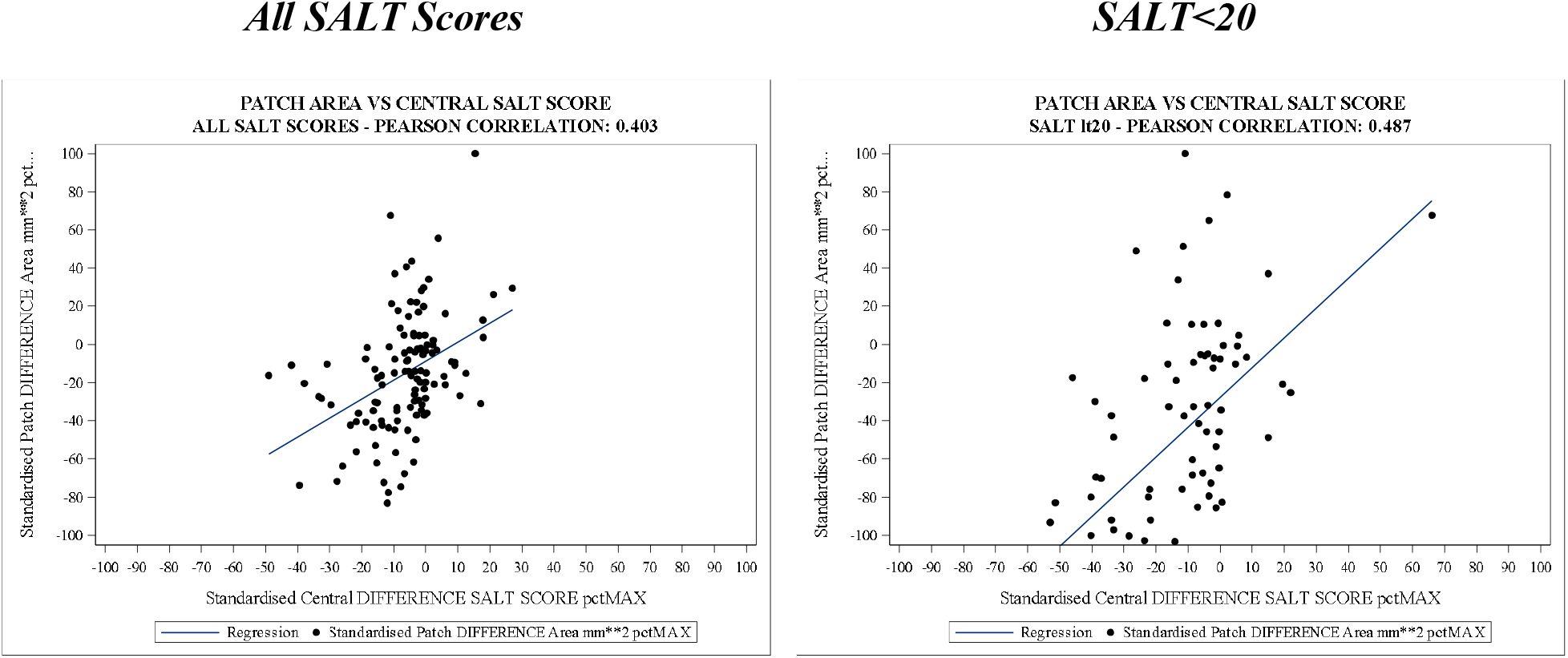
Correlation Central Rating vs Patch Area Measurement.

### Central vs local rating: Reproducibility (comparison)

Overall, SALT score comparisons for Central vs Local rating were more variable with much less Agreement: measurement error=16.2, repeatability coefficient=31.7 and a lower ICC=0.54 (Figure 4A).

**Figure 4A:**
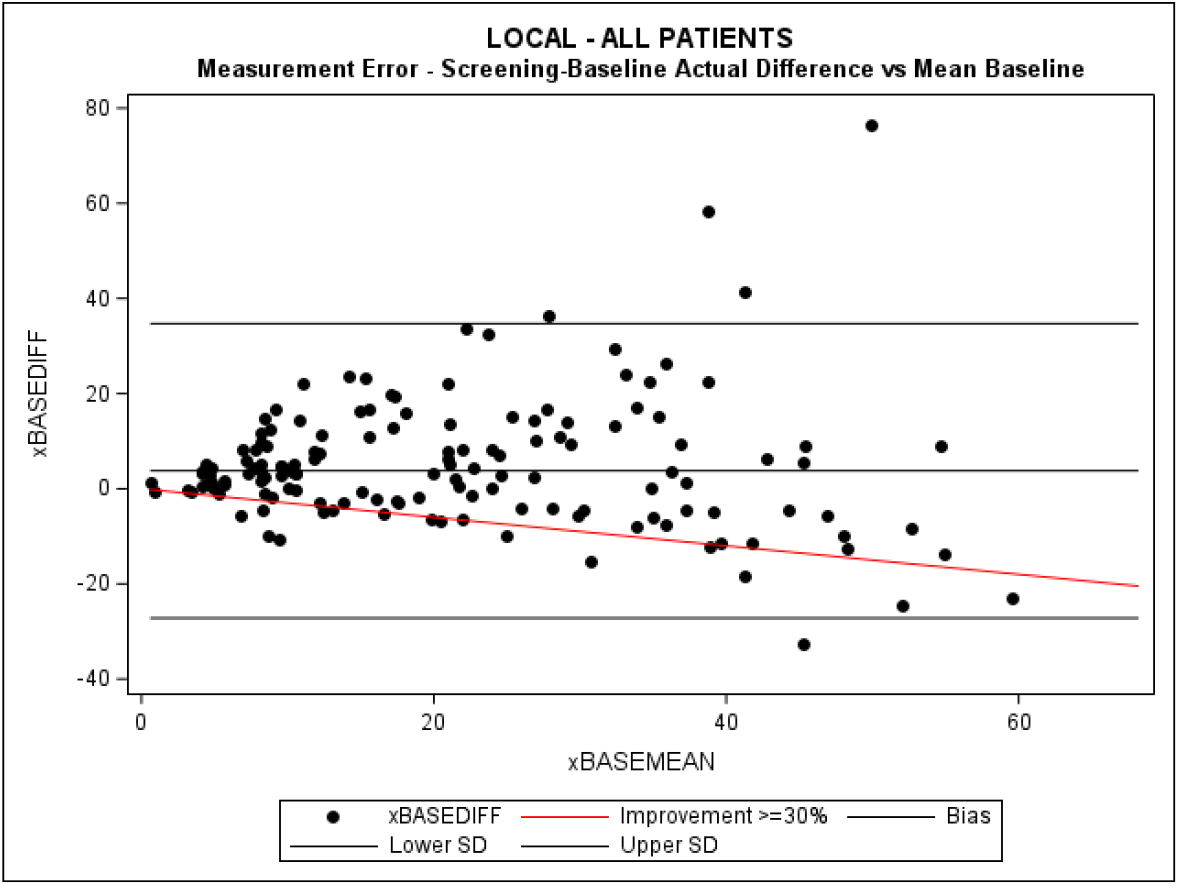
Reproducibility (Comparison) – Local vs Central Baseline.

Agreement between the two methods was also limited with high LoAs around +/-30 over the entire SALT range. In addition, the variability of differences appeared to increase considerably with higher SALT scores, and again there appeared more differences in scoring above SALT 20.

In addition, the overall mean difference, +3.76, indicated potential bias between the two methods where Local raters consistently scored higher in comparison to Central rating and potentially therefore over-estimated the amount of hair loss. Therefore, the Local rating scoring was overall assigning worse hair loss at Baseline, although as shown below this was skewed depending on the end of the scale. However, overall, this bias did not occur with Central rating.

Separate plots for SALT <20 and ≥20 (Figure 4B) suggest LoAs of 20 and 42, respectively, suggesting better, albeit still limited accuracy, with the lower the SALT scores and with bias between the two methods still evident. In comparison the Central rating LoA was 5 with SALT<20.

**Figure 4B:**
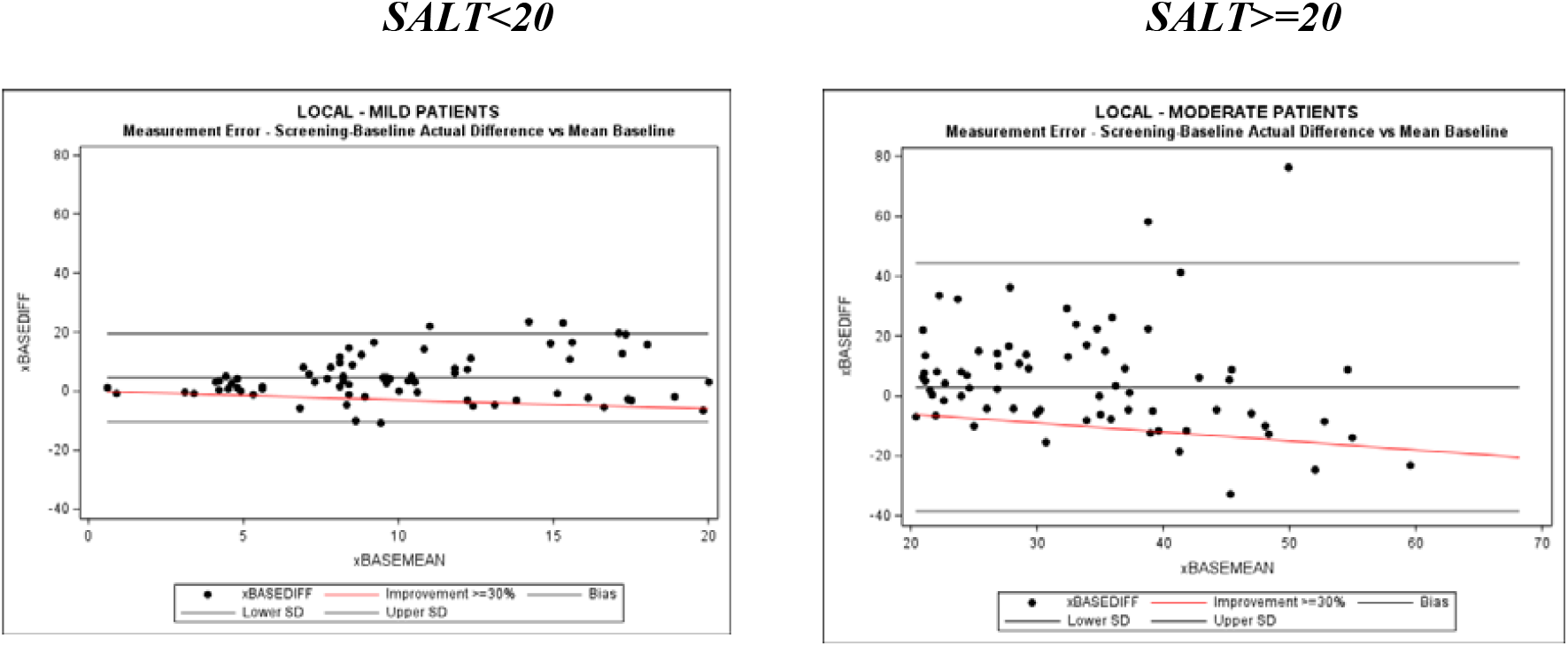
Method Comparison (Reproducibility) – Local vs Central Baseline.

Correlation plots (Figure 4C) for the two methods also demonstrate less agreement when compared against the line of uniformity (black line). Of note, the higher the SALT score the more data points lie below the line of equality, and the lower the SALT score the more data points lie above the same line. This indicates that the differences between the Local and Central rating methods are more pronounced at the extremes of the scale i.e local raters score subjects higher the larger the SALT score and lower the smaller the SALT score. As this is occurring prior to randomisation, it maybe that allowance is being made for the requirements of the protocol and this influences the assigned score.

**Figure 4C:**
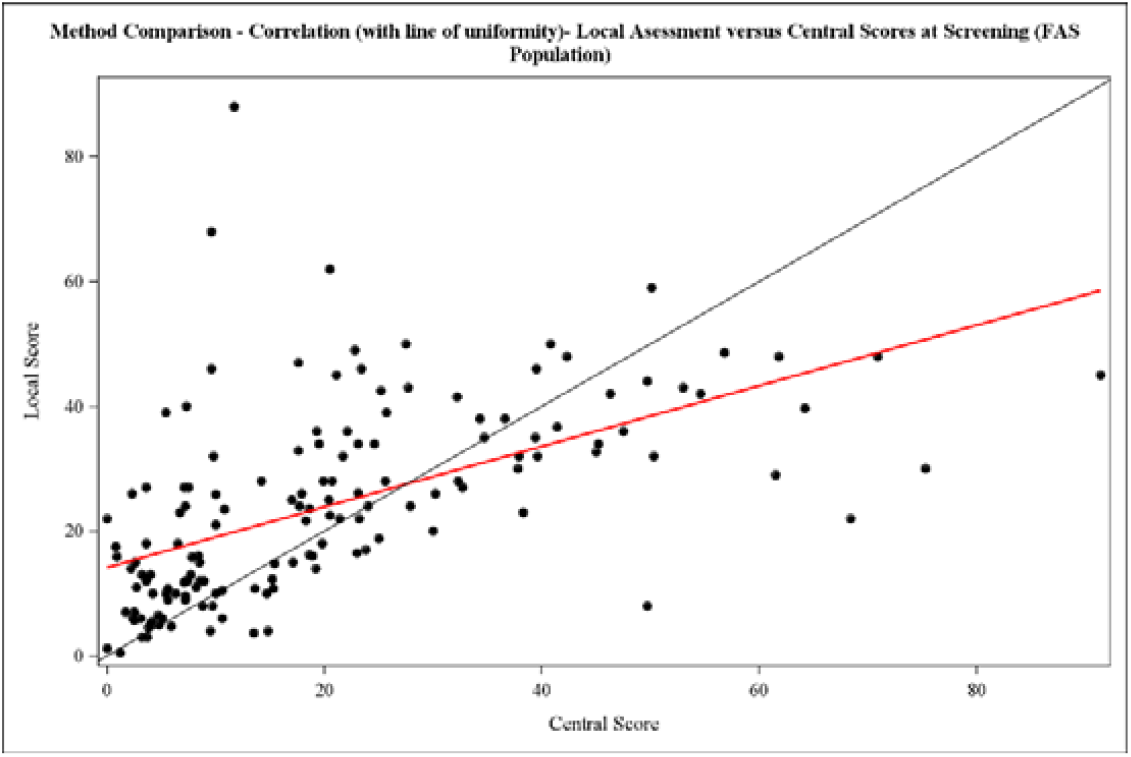
Method Comparison (Reproducibility) – Measurement Bias: Central vs Local. NB: Regression line in Red, Line of uniformity in black

The bias between the two measurement methods was assessed using a one-sample t-test. The actual overall mean difference of 3.76 (SE 1.27) showed a statistically significant difference (p=0.0035) confirming the bias observed between Central and Local rating.

### Impact on Phase 2 clinical trial outcome

The simulations performed demonstrate that substituting Local rating assessments for Central scoring would result in the likelihood of statistically significant outcomes of between 13% to 43% (power) depending on the endpoint selected. At best then, the likelihood of success would be reduced by half, as a result of inflating variability generated with Local raters (Table 1).

**Table 1:** Phase 2-Local Score Model Simulation of Trial Outcome – N 8,000-10,000 trials.

| Measure | Statistically Significant Outcome | N Trials | Power (%)* |
| --- | --- | --- | --- |
| Actual Mean Change from Baseline | No | 7397 |  |
|  | Yes | 2079 | 21.9 |
| Percentage Mean Change from Baseline | No | 8288 |  |
|  | Yes | 1222 | 12.8 |
| Responder Rate ($\geq 30\%$ Improvement) | No | 5173 | |
|  | Yes | 3869 | 42.8 |
| Responder Rate (100% Improvement) | No | 6498 |  |
|  | Yes | 1850 | 22.2 |

\*NB: Power is proportion of 'Yes' outcomes (Type II error is 'No' proportion when outcome correct)

## Discussion

The results consistently show that Central rating provides an accurate representation of the quantity of hair loss, which minimises the error in SALT scores by removing the between rater variability and providing consistency in measurement. It also clearly demonstrates that there is an issue with accuracy with Local rating, notably at the lower end of the SALT score range and this is in agreement with other publications^7^,where as a result Local rating has been considered unwise.

Concern with Central rating appears to be centred around belief that photographs will not completely reveal areas of hair loss and so the SALT assessment will be under-scored. In the situation of a controlled, double blind, randomised clinical trial, statistically this will be equally likely to occur in both treated and control groups and therefore the magnitude of the derived treatment effect will be unaffected and unbiased. In addition, clinically one could also argue that if done ‘once’ this is likely to be done at ‘all’ subsequent visits and so would not affect the change in score. This would have to be undertaken ‘wrongly’ with treatment during later assessments to overestimate the treatment response. Conversely, with a placebo control this would have to be performed in a ‘visa versa’ fashion to overestimate the treatment – control difference. Ultimately, this would suggest a specific observer or systematic bias, which would only be possible in a local rating setting where ordering is known. Again, this would justify the requirement for Central rating as a ‘fairer’ comparison.

Our own experience suggests ‘under’ scoring is only a risk in the rarer cases of diffuse hair loss (actually an exclusion criteria in our Phase 2 trial), rather than the more commonly seen patchy appearance and here clear instructions are provided to photographers to ensure hair is ‘clipped’ accordingly to reveal the patch. In this situation, supporting software with photography allows aligning of grids for more accurate scoring which is not possible with the naked eye in Local scoring. This is also supported by a research team which developed a neural network program to support precision in image scoring.6 Ultimately these situations justify and underpin the basic premise of the randomised, double-blind, placebo controlled, clinical trial design originally advocated by Austin Bradford Hill. Randomisation allows both known and unknown/uncontrollable influences to be equally distributed between the treatment groups, ensuring statistical validity of the estimated treatment effect.

The basic question still to resolve is - How is it possible to ensure accurate assessment of treatments in clinical trials involving all Alopecia areata patients? So far, we have highlighted that, although it is pretty much accepted that SALT scoring the amount of hair loss is appropriate, the actual method of scoring is much more problematical. A recent clinical paper highlights the practical advantages of using photography with images taken according to standardised protocols and equipment with training for Nursing staff and Patient alike to ensure ‘superior assessment’ of the progression of hair loss and treatment efficacy in clinical trials.^12^ Although ‘challenges’ were noted including: Standardising patient activity (hair styling/cutting, dyeing cycles), type of patient (fair hair, fair skin for contrasting) positioning (for diffuse hair loss, widespread small patches, longer hair, curly hair, hair extensions), the conclusion was that the benefits of objective assessment in serial measurements outweighed the risks with accurate and reproducible findings.

In mild-moderate patients Local rating certainly seems ill advised due to problems of measurement accuracy. Whilst even in severe patients where measurement accuracy is of less importance there is still considerable concern for variability and clinical judgement on the classification of improvement. A good example of this is seen in severe cases where improvements were graded according to a classification system of SALT scores (Investigator’s Global Assessment: AA-IGA) to reduce the inter-rater error and to provide a more clinically relevant outcome measure.^13^

Our pragmatic advice would be that application of numeric accuracy is most relevant to a Phase 2 setting where precise determination of the magnitude of the treatment effects are paramount in the ‘learning’ environment of early phase development. Subsequently, following progress to ‘confirmation’ of efficacy in the much larger generalised Phase 3 setting, crucially more meaningful clinical outcome results other than just a numerical score for improvement become necessary. As such, clinical judgement of what is a meaningful outcome following treatment is essential. In this respect, development of the AA-IGA suggests that there is still need for clinical consensus of what is a relevant clinical outcome. In mild to moderate patients a clear benchmark is needed and we would support a ‘complete regrowth’ endpoint with the subject able to satisfactorily stop treatment. This would involve a locally rated SALT score 0 with clear support with clinical signs of restored terminal hair growth as this then satisfies the lowest common denominator for agreement between clinicians, and would seem to be the only possible real practical solution given numerical issues. As a footnote, it is worth noting that we haven’t mentioned the potential need to consider the influence of patient reported outcomes in the definition of successful response. However, health related quality of life (HRQoL) measures are proving equally difficult to assess in the AA patient group and we have investigated this further in a separate article.

In conclusion, this study using numerical assessments, central rating was found to be the most appropriate in a Phase 2 ‘learning’ stage of clinical development, providing an accurate representation of the quantity of hair loss whilst minimising the error in SALT scores and providing consistency in measurements.

## Data Availability

will be made available on request

## Notes

### Competing Interest Statement

The authors have declared no competing interest.

### Clinical Trial

NCT06402630/ Eudract 2021-004145-20

### Funding Statement

Funding from Soterios LTD

### Author Declarations

London South West Ethics

